# ELIPSE-COL: A novel ELISA test based on rational envisioned synthetic peptides for detection of SARS-CoV-2 infection in Colombia

**DOI:** 10.1101/2020.11.13.20230060

**Authors:** Adriana Arévalo, Carlos Franco-Muñoz, Sofía Duque-Beltrán, Lyda Muñoz-Galindo, María Herrera-Sepulveda, José Manuel Lozano, Luz Mary Salazar, Martha L. Ospina-Martinez, Marcela Mercado-Reyes

## Abstract

**Background:** COVID-19 pandemic caused by infection with the betacoronavirus SARS-CoV-2 is the greatest public health defiant on a global scale in the last 100 years. Governments and health Institutes face challenges during the pandemic, related to the diagnosis, mitigation, treatment, and timely detection after the epidemic peak for the prevention of new infections and the evaluation of the real impact of the COVID-19 disease in different geographic areas. To develop a valuable tool to study the seroprevalence of SARS-CoV-2 infection in Colombia, an “*in-house*” ELISA was achieved for the detection of IgG anti-SARS-CoV-2 antibodies in serum.

**Methods:** The test was standardized using an antigenic epitope “Pool” of the synthetic peptide as antigen derived from antigenic regions of the spike, nucleocapsid, envelope, and membrane structural proteins, which were designed, based on the genomic information of SARS-CoV-2 circulating in Colombia. In the ELISA standardization process, 94 positive sera were used, including sera from asymptomatic and symptomatic patients (mild and severe) and 123 negative sera, including pre-pandemic historical negatives originating from patients living in arbovirus endemic areas or patients with a history of respiratory diseases and sera from patients with a negative rRT-PCR test for SARS-CoV-2.

**Results:** The in-house peptide ELIPSE-COL test showed promising performance, being able to detect reactivity in sera from asymptomatic and symptomatic patients. The sensitivity and specificity of the assay were 91.4% and 83.7% respectively.

**Conclusion:** ELIPSE-COL assay was developed as an ELISA test using synthetic peptides for the study of the seroprevalence of SARS-CoV-2 infection in Colombia.

**SUMMARY BOX:**

- Detection of IgG anti-SARS-CoV-2 antibodies is required for the evaluation of the pandemic impact and vaccination strategies.
- ELIPSE-COL is an in-house test based on synthetic peptides as antigen derived from antigenic regions of the spike, nucleocapsid, envelope, and membrane structural proteins.
- The sensitivity and specificity of the assay were 91.4% and 83.7% respectively suggesting a promising performance.
- ELIPSE-COL test is a valuable tool for the study of seroprevalence in Colombia.

## INTRODUCTION

The life-threatening respiratory illness called COVID-19 caused by the SARS-CoV-2 betacoronavirus emerged in December 2019 in Wuhan (China) and impacted the public health on a global scale with no precedence in the 21st century (1, 2). The COVID-19 disease caused almost 119.000 cases and 5.500 deaths in China, however, after global dissemination and over a year of the pandemic, the SARS-CoV-2 infection has cost over three million of life lost and near 182 million cases (3). The first case of COVID-19 in Colombia was reported near to four millions of cases and 110 thousand deaths to Jul 2021(4).

The genomic information of SARS-CoV-2 is encoded in a single, positive-stranded RNA (ssRNA[+]), with near 30.000 nucleotides long. The first of the nine open reading frames (ORF) composing the SARS-CoV-2 genome is subdivided in ORF1a and ORF1b by ribosomal frameshifting and encodes two polyproteins which are processed into non-structural proteins involved in subgenomic/genome length RNA synthesis and virus replication. In the subgenomic region, the structural proteins, Spike (S), Envelope (E), Membrane (M), and Nucleocapsid (N) are encoded in subgenomic mRNA transcripts within ORFs 2, 4, 5, and 9, respectively (5). The Structural proteins of SARS-CoV-2 have been exploited as an ideal antigen from different applications including, IgG or IgM chromatographic rapid test (6), ELISA test (7) and to produce polyclonal and monoclonal antibodies in animal models (8). Most of the reported applications use recombinant DNA approaches, however, synthetic peptide technologies are also available as desirable ligands in the development of new antibody detection tests.

Short peptides became important reagents in epitope mapping since the technology to synthesize large numbers of peptides on solid support was developed in the 1980s (9, 10). Peptides can be synthesized in large quantities with very high purity and without recombinant cellular expression systems, and those who specifically bind to target antibodies are valuable ligands useful in serological assays (10, 11). Relative chemical stability of short peptides and the relative ease with which they can be synthesized and manipulated (12, 13).

According to the epidemiological surveillance of the COVID-19 pandemic in Colombia, the country was passed through at least three epidemic peaks, the last one with a devastating increase in dead count. The COVID-19 vaccination program starts at February 2021 and at this moment timely case detection, strategies for the prevention of new infections, and the evaluation of the true impact of the COVID-19 disease in Colombia are needed. Taking all this into consideration, this study aimed to develop an “in-house” ELISA test using rational designed synthetic peptides as an antigen for the detection of total anti-SARS-CoV-2 antibodies in sera from humans being infected with SARS-CoV-2. The achieved test is ELIPSE-COL, a valuable tool for the study of SARS-CoV-2 infection in Colombia.

## METHODS

### Ethical considerations

This work was approved by the Ethics and Research Methodologies Committee (CEMIN) of the National Institute of Health (created by resolution 395 of April 4, 2017) who determined that the work meets the technical and ethical requirements for which they granted ethical approval registered in Act No. 7 of April 13, 2020.This work was developed according to the national law 9/1979, decrees 786/1990 and 2323/2006, which establishes that the Instituto Nacional de Salud (INS) from Colombia is the reference laboratory and health authority of the national network of laboratories and in cases of a public health emergency or those in which scientific research for public health purposes as required. This study was performed following the ethical standards of the Declaration of Helsinki 1964 and its later amendments. The information used for this study comes from secondary sources of data that were previously anonymized and do protect patient data. All necessary patient/participant consent has been obtained and the appropriate institutional forms have been archived.

### Peptide Design

Worldwide and Colombian SARS-CoV-2 genome sequences were retrieved from GISAID and NCBI databases (https://www.gisaid.org/, https://www.ncbi.nlm.nih.gov/sars-cov-2/). Substitution matrices of nucleotides and amino acids of structural proteins were generated from a multiple sequence alignment with the SARS-CoV-2 genomes using the Muscle Algorithm (14) in MEGA X (15). *In silico* analyses were performed to identify the presence of LB epitopes, including the presence of proteasome cleavage sequence sites, HLA-I and HLA-II different length binding epitope sequences regarding endosomal and phagosome-lysosome protease cleavage sites in each ORF coded sequence using LBtope (http://crdd.osdd.net/raghava/lbtope/), ABCpred main page http://crdd.osdd.net/raghava/abcpred/ and BEpiPred-2.0 (http://www.cbs.dtu.dk/services/BepiPred/) bearing a threshold: 0,5. Aminoacid sequences containing antigenic regions being exposed in a non-conformational configuration were selected for peptide design.

Optimization of peptide designs was performed using the peptide analyzing tool of ThermoFisher (https://www.thermofisher.com/co/en/home/life-science/protein-biology/peptides-proteins/custom-peptide-synthesis-services/peptide-analyzing-tool.html) to calculate the peptide physical-chemical properties, including the charge-pH map, isoelectric point (pI), hydrophobicity, and mass and predict the ease of peptide synthesis and purification.

### Solid-phase Peptide Synthesis

The peptides designed were synthesized from two sources: the first one was a commercial ThermoFisher platform Peptide Synthesis Services, and the second one was produced in collaboration of two research groups (National Health Insitute of Colombia and National University of Colombia). Therefore, the selected peptides were synthesized manually using a solid-phase synthesis by 9‐fluorenylmethoxycarbonyl (Fmoc) strategy. Solvents and soluble reagents were removed by filtration. Washings between deprotection, couplings, and subsequent deprotection steps were carried-out with *N,N*′‐dimethylformamide (DMF) (5 x 1min), dichloromethane (DCM) (4 x 1min), Isopropyl alcohol (IPA) (2 x 1min) and DCM (2 x 1min) using 1.5 mL of solvent/50mg of resin each time. The Fmoc group was removed from the resin by two treatments of 15 min with piperidine‐DMF (25:75 v/v). Couplings were performed at 20°C and monitored using standard Kaiser tests for solid‐phase synthesis. after Fmoc removal of the commercially available Rink amide resin (50 mg, 0.46 mmol/g), a first Fmoc-amino-acid (0.115mmol, 5.0 equiv.) was added with 1-hydroxy benzotriazole (HOBt) (18.2 mg, 0.115 mmol; 5.0 equiv.) and *N,N*′-dicyclohexylcarbodiimide (DCC) (23.7 mg; 5.0 equiv.) as coupling reagents dissolved in DMF/DCM (7:3, v/v) and the coupling reaction was stirred for 2 hours. Next, the Fmoc group was removed, and a second Fmoc-amino-acid was incorporated in the resin using the same conditions. The Fmoc removal and the coupling reactions of the rest of the Fmoc-amino-acids were carried out under the same conditions using 5 equiv./coupling. Finally, monomer peptide was Fmoc deprotected and cleaved from the resin by treatment with a mixture of trifluoroacetic acid-water-triisopropylsilane (TFA/H2O/TIS) (95.0:2.5:2.5) for 6 hours followed by filtration and precipitation with cold diethyl ether (Et_2_O). Crude products were then triturated 3 times with cold Et_2_O, dissolved in the system water-acetonitrile (H_2_O:CH_3_CN) (9:1 v/v), and then lyophilized (9). Protocols adapted from published data were also used (16).

### Samples

Ninety four sera from patients diagnosed with COVID-19 by the confirmation of SARS-CoV-2 infection by rRT-PCR were selected. Fifty six of them were from asymptomatic patients and thirty eight sera from patients with symptoms between mild and severe symptoms. Blood samples were taken from each of the asymptomatic and symptomatic patients between 14 to 21 days after the first date of diagnosis by rRT-PCR. One-hundred-twenty-tree negative samples were selected from the Biological samples Bank of the National Institute of Health of Colombia and which were corroborated by rRT-PCR of the absence of SARS-CoV-2. Sixteen of them came from endemic areas for arbovirosis (Zika. Dengue, Chikungunya) and which were obtained months before the first case of COVID-19 was reported in Colombia. Two hundred seventeen samples were included in total.

### ELIPSE-COL: immunodiagnostic assay for the detection of anti-SARS-CoV-2 antibodies

Polystyrene 96-flat bottom plates (Inmulon IB) were immobilized overnight with a “Pool” of 7 proteins peptides: 2 of Spike protein peptides, 3 Nucleocapsid proteins peptides from different antigenic regions, 1 Membrane protein-peptide, and 1 Envelope protein-peptide all of them derived from antigenic regions of SARS-CoV-2. The “Pool” of proteins peptides (antigen) was adsorbed to the polystyrene plates in concentrations ranging from 5 µg/ mL to 40 ug/ mL in a carbonate/bicarbonate buffer at a pH of 9.6 at 4°C. Followed three washes with PBS using a Well Washing 4MK2 machine (Thermo Scientific).

Non-specific binding was blocked with a solution of 1-5% of skimmed-milk in 0,15M (phosphate-buffered solution) PBS-0.05% (v/v) tween-20. Followed three washes with PBS using a Well Washing 4MK2 machine (Thermo Scientific).

Different dilutions from 1:25 to 1:200 of human sera samples were poured onto peptide adsorbed-ELISA plate wells and incubated for 3 hours at 37°C. Followed three washes with PBS using a Well Washing 4MK2 machine (Thermo Scientific). Then a goat alkaline phosphatase anti-human-Ig-conjugate was poured at 1:500 of dilution on PBS-Tween-20 and incubated for one hour at 37°C to bind specific human antibodies to SARS-CoV-2 epitopes. After performing standard washings, the test was developed with a 1 mg/mL p-nitrophenylphosphate solution in 0,1M diethanolamine, pH 9.8 to reveal those antibodies specifically binding to designed virus epitopes by a yellow color appearance which was detected on a microplate reader (Multiscan EX®, ThermoFisher scientific,) at 405 nm. When necessary enzyme activity was stopped by adding a 3N NaOH solution before absorbance reading.

### Statistical analysis

#### Determination of the cut-off point

The cut-off value was determined taking into account the absorbance value that differentiated between positive and negative samples. This value was calculated by adding two standard deviations to the mean value of the negative samples, which offers 95% reliability (17, 18).

#### Coefficient of variation (CV)

The coefficient of variation (CV) was obtained by intra-plate repeatability and inter-plate repeatability. To evaluate intra-plate repeatability, four replicates of 24 samples were tested. Inter-plate repeatability was evaluated by using the same samples in four different plates. CV was calculated as CV = (standard deviation/mean) × 100% Agreement within CIEP and peptide ELISA data was obtained by analysis of the same serum (19).

#### Diagnostic discrimination

The following diagnostic indicators were deduced by the given formulae:

- Sensitivity (SE): Number of true positive samples=true positive sample number number of false-negative samples
- Specificity (SP): Number of true negative samples=number of true negative samples number of false-positive samples
- Positive Predictive Value (PPV): Number of true positives / The sum of true positives and false positives
- Negative Predictive Value (NPV): Number of true negatives / The sum of true negatives and false negatives
- Positive Likelihood Ratio (CPP): Sensitivity / 1 -Specificity
- Negative Probability Ratio (NPC): (1 -Sensitivity) / Specificity
- Coincidence rate (CR): true positive sample number + true negative sample number / number of total samples

#### Descriptive Statistical Analysis

The average value of the absorbance values (three absorbance values obtained per sample) was calculated for both positive and negative samples. As well as the dispersion means of the mean absorbance values (Standard Deviation). A comparison of mean ranges between the groups was performed using the Mann-Whitney U test and the Kruskal-Wallis one-way analysis of variance followed by Dunn’s multiple comparison test as required. All statistical tests were performed with a 5% level of significance. Graphs were constructed in GraphPad Prism software version 7.0.

## RESULTS

### Antigenic peptides

A representative antigenic peptides were pooled and used as the antigen for ELIPSE-COL and following named “Pool”. Such selective mix antigenic peptide representative for SARS-CoV-2 was achieved by combining the most representative synthesized peptide epitopes containing antigenic regions of the structural proteins: ThermoFisher platform Peptide Synthesis Services (1 antigenic region of the Spike protein, 2 antigenic regions of the nucleocapsid protein) and those designed by the research groups, including antigenic regions correspond to a region of the Spike protein, 1 region of the nucleocapsid protein, 1 region of the envelope protein, and 1 region of the membrane protein.

### ELIPSE-COL: ELISA test developed for serodiagnosis of SARS-CoV-2 infection

ELIPSE-COL immunodiagnostic test was developed taking into account the characteristics hydrophobic properties of the peptides used (“Pool of Peptides”) as antigens. The peptides adhered to Immulon IB polystyrene plates once the physicochemical characteristics of Immulon IB had been analyzed versus polypropylene, polycarbonate, and nylon, also considering the importance of using plates transparent flat bottom for adequate passage of light at a wavelength of 405 nanometers (nm) through each well. Sodium carbonate/ bicarbonate buffer solution pH 9.4 at 0.2 M was selected as a coating solution and adherence of peptides to plate wells. This pH 9.4 value helps the solubility of many proteins and peptides by ensuring that most proteins are not protonated with an overall negative charge, which aids in the adsorption of peptides to the positively charged plate, others solutions such as Tris Buffer Saline (TBS) or Phosphate Buffer Saline (PBS) were considered (20, 21). Coated wells with the “Peptide Pool” could leave free hydrophobic/hydrophilic sites on the polystyrene bottom of the plate. These sites must be blocked with a specific step called “post-coating” to avoid non-specific binding of subsequent reagents. In the ELIPSE-COL test, the 5% skim-milk solution was used as a viable option for blockade considering its commercial value and affordability, although 5% of serum albumin could also be used. Both allow a real blockade in such a way that it is possible to differentiate between positive and negative serum (Fig. 1). Antigen-antibody binding can be visualized using alkaline phosphatase-linked conjugate. Unlike peroxidase, alkaline phosphatase activity is not altered by exposure to antibiotics and other agents such as thimerosal or sodium azide. This means that the enzyme can be stored for a defined time, even in a non-sterile environment (22). Taking these characteristics into account, a test using alkaline phosphatase can be performed in any laboratory, with minimal conditions without affecting the results.

**Figure 1.**
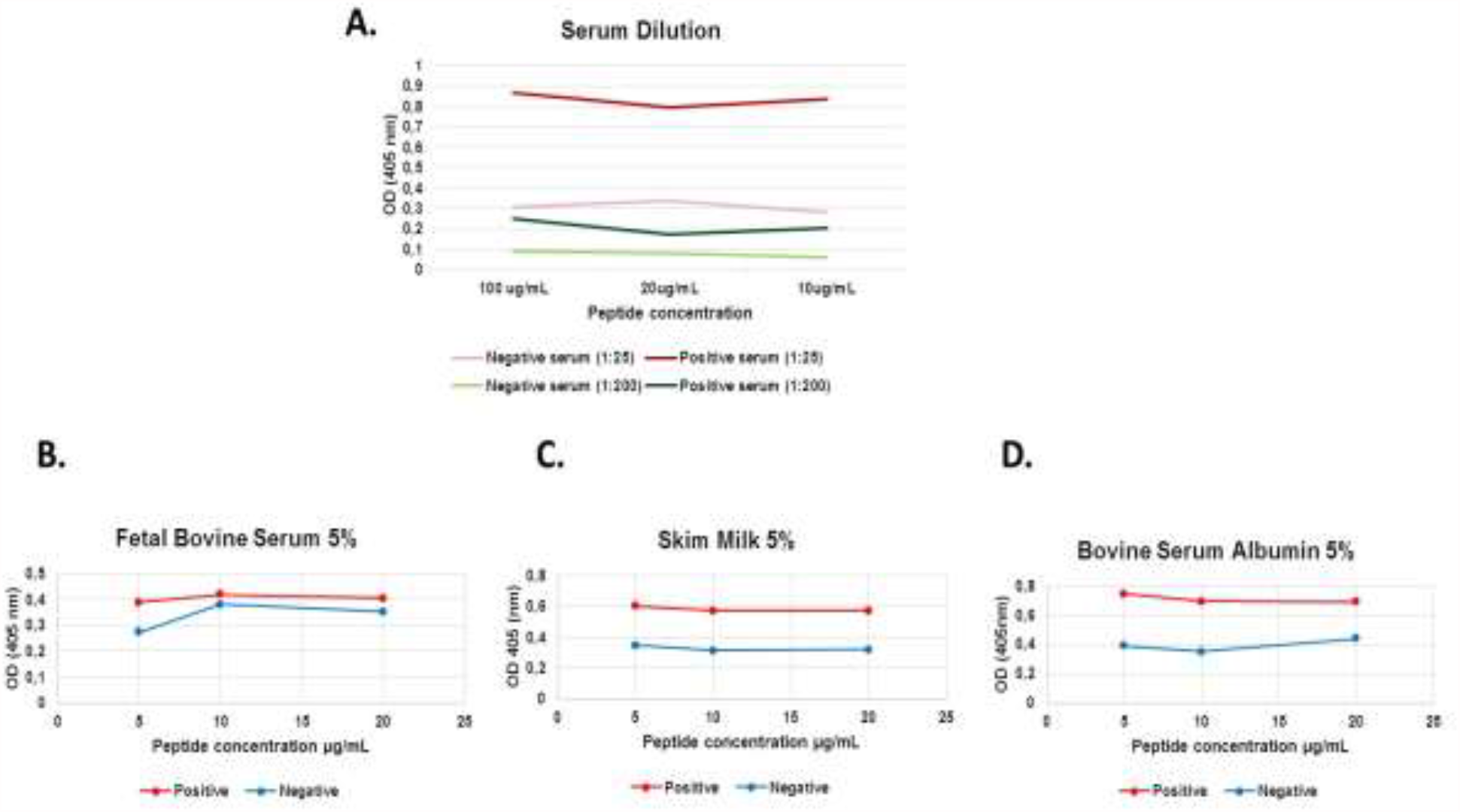
ELISA test developed for serodiagnosis of SARS-CoV-2 infection:. **A**. Optimal serum dilution evaluation. **B**. Fetal bovine serum at 5% as the blocking solution. **C**. Skim milk at 5% as the blocking solution. **D**. Bovine serum albumin at 5% as the blocking solution.

After the standardization process, it was found that the optimal test conditions corresponded to 25 µg/ml of the peptide pool, serum dilution of 1:25 with incubation of 3 hours, and dilution of 1: 500 of the peroxidase-conjugated anti-human IgG antibody (Fig. 1).

The coefficient of variation of the test in intra-plate repeatability and inter-plate repeatability experiments had a dispersion from 5.4% to 19.25%, with an average variation of 13%. It has been reported that the CV for this parameter should not exceed 20%, being optimal those below 5% and 10% respectively.

### Diagnostic discrimination

The ELIPSE-COL test based in synthetic peptides shows a differential serological reaction to the COVID-19 positives and negatives sera. The variation in the amplitude of the standard deviation bars in the group of sera from positive patients (with COVID-19) (Fig. 2) can be attributed to the inclusion of sera from convalescent patients (Hospitalized) for which values of absorbance up to 1.7 were registered, unlike the values observed in the group of sera from patients without COVID-19. Statistically significant values were found between the median absorbances of sera from patients with COVID-19 versus sera from patients without COVID-19.

**Figure 2.**
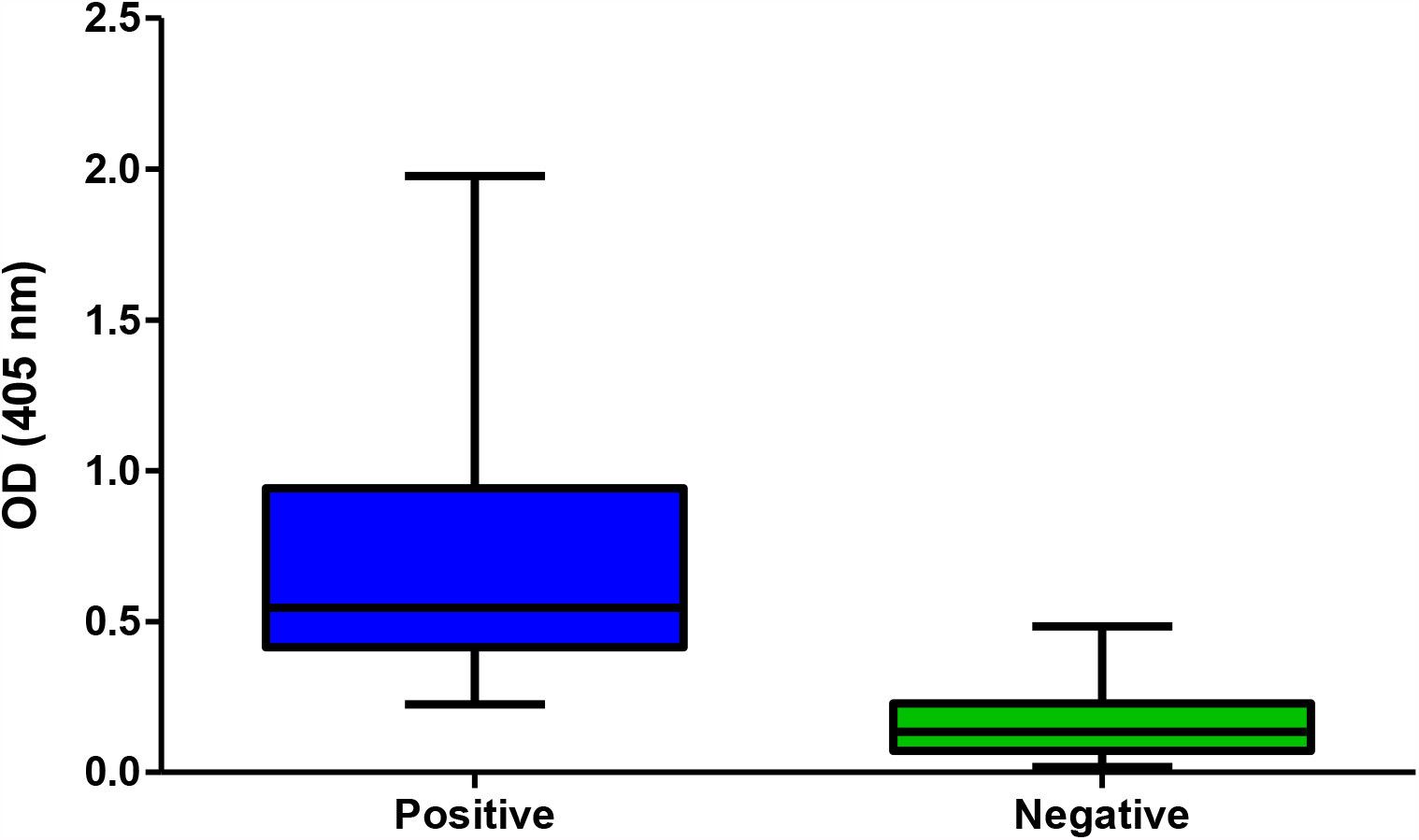
Serological reaction to synthetic peptide-epitopes of antibody-serum from individuals being SARS-CoV-2 positive and negative RT-PCR tested.

The absorbance cut-off value was estimated at 0.285 and it was applied to differentiate between positive and negative samples. Diagnostic classification of the study groups for the ELIPSE-COL test is shown in table 1, 86/94 positive sera, and 103/123 negative sera were properly classified by the “*in-house”* test.

**Table 1.** Diagnostic classification of the study groups by the ELIPSE-COL test

|  | rRT-PCR<br>(+) | rRT-PCR<br>(-) | Total |
| --- | --- | --- | --- |
| **ELIPSE-COL**<br>**(+)**<br>**OD ≥ 0,278 cut-off** | 86 | 103 | 106 |
| **ELIPSE-COL**<br>**(-)**<br>**OD 0,278 cut-off** | 8 | 20 | 111 |
| **Total** | **94** | **123** | **217** |

The diagnostic classification by positive/negative groups obtained for the test was used to calculate the parameters of diagnostic accuracy and operational performance of the ELIPSE-COL immunodiagnostic test, which showed the following values for each of the indicators (Table 2):

**Table 2.** Diagnostic accuracy and operational performance ELIPSE-COL test

| Indicator | Value | Confidence intervals (CI 95% ) |
| --- | --- | --- |
| Sensitivity (SE): | 91.49% | (IC 95% 83.92% - 96.25%) |
| Specificity (SP): | 83,7% | (IC 95% 76.01% - 89.78%) |
| Positive Predictive Value (PPV): | 83.7% | (IC 95% 74.13% - 86.58%) |
| Negative Predictive Value (NPV): | 95.3% | (IC 95% 86.85% - 96.17%) |
| Accuracy (CR): | 90.7% | (IC 95% 86.04% - 94.22%) |
| Median absorbance values of sera from COVID-19 patients | 0.803 |  |
| Median absorbance values of sera from patients without COVID-19: | 0.104 |  |
| Mann Whitney test "p" value: | <0.0001 |  |

## DISCUSSION

The detection of anti-SARS-CoV-2 IgG antibodies during the COVID-19 pandemic has been focused on using immunodiagnostic tests that use recombinant proteins as capture antigen. (23-25). These immunodiagnostic tests differ in their sensitivity and specificity values, which makes it difficult to use a single one for the detection of anti-SARS-CoV-2 antibodies worldwide. During the pandemic, the ELISA tests more often used are based on recombinant protein (25) designed with the first SARS-CoV-2 genomes, this proteins could not recall the changes in the genomic diversity of SARS-CoV-2 that appears after the emerging of the virus and its global dissemination (26).

The “pool” of synthetic peptides were designed based on the genomic information of SARS-CoV-2 (Coronavirus) circulating in Colombia integrate the most important antigenic regions of the structural proteins of the coronavirus, which is a guarantee to detect antibodies targeted against different parts of the virus, which favors detection thus, non-synonymous substitutions occur in the virus genome. In fact, in the design of the peptides, it was taken into account that 12 lineages of SARS-CoV-2 circulate in Colombia, which present changes in the sequences concerning the coronavirus initially characterized in Wuhan (27).

One of the most studied amino acid changes in SARS-CoV-2 is the substitution of aspartic acid for glycine at position 614 of the S1 subunit of the Spike protein (28) and it is known that this substitution interrupts an antigenic region in this protein (29). Given the above, if this were to happen at one point in time and which could occur, it would not drastically affect the performance of the detection of anti-SARS-CoV-2 antibodies due to the presence in the peptide pool of the other proteins that comprise it and that were developed conserving 2 different peptides of the Spike protein, 3 different peptides of the Nucelocapsid, 1 peptide of the Envelope and 1 peptide of the Membrane that is also antigenic and are used in ELIPSE-COL developed in the present study

Several of the immunoassays developed for the detection of anti-SARS-CoV-2 antibodies tend to focus exclusively on the Spike protein and particularly on the receptor-binding domain (RBD) of the protein as an antigen to detect neutralizing antibodies given the SARS-COV-2 pathway via ACE2 receptor (30, 31). However, this is beyond the scope of an ELISA test and the presence of this type of antibody must be verified with a plaque reduction neutralization test (PRNT) (7, 32), therefore, this guidance may limit the performance of an ELISA test that is designed to detect whether the person is or has been exposed to SARS-CoV-2. Nonetheless, some studies have been suggested that epitopes on the SARS-CoV-2 spike protein could be correlated with the presence of neutralizing antibodies in COVID-19 patients (33, 34) or with the disease severity (35).

One of the disadvantages of using peptides is their non-renewable after synthesis, unlike recombinant antigens that can be produced in cell lines at different times. However, As in the case of this study, the production of peptides can be obtained individually at a scale of 100 milligrams, and in the immunodiagnostic test developed (ELIPSE-Col) only 6 micrograms of the “peptide pool” are used per test. Given the above, a production of 100 milligrams with an approximate time of eight days allows the testing of anti-SARS-CoV-2 antibodies in 16,600 people, which is not a critical disadvantage given its scale of production obtained in the study. Furthermore, if a substitution in the SARS-CoV-2 sequence were to occur, the immunodiagnostic test developed with them could be adaptable in a week and this would have a positive impact on preserving the performance and functionality of ELIPSE-Col. An advantage of synthetic peptides could be to produce them to same or bigger scale to describe before at the time the SARS-CoV-2 change.

The synthetic peptides used in the ELIPSE-COL immunodiagnostic test are an advantage in the performance of the test due to their specificity, which avoids cross-reactions with endemic arboviruses (36) and other coronaviruses such as (Human Coronavirus 229E, Human Coronavirus NL63, Human Coronavirus OC43, and Human Coronavirus HKU1 circulating in Colombia (37).

As far as the scientific literature allows, immunodiagnostic tests to detect SARS-CoV-2 antibodies using synthetic peptides as performed. Meng and co-workers used a “pool” of peptides from the Spike protein to detect immunodominant regions of anti-SARS-CoV-2 IgG (33) and the ELIPSE-COL used a “Pool” of peptides from different regions of SARS-CoV-2 but with the purpose of detect anti-SARS-CoV-2 antibodies in patient serum.

The use of peptides for immunodiagnosis in the case of SARS and MERS is known when Woo, et. al., in 2005 developed an ELISA test to detect anti-SARS-CoV IgG and IgM antibodies using peptide pool as antigen independently of the Spike and Nuclocapsid proteins with sensitivities of 74.7% and 94.75%, respectively (38). The presence of both targets (Spike and Nucleocapsid) and other structural peptides of SARS-CoV-2 in the ELIPSE-Col “Pool” its performance with a sensitivity of 91,4%.

In the current SARS-CoV-2 Pandemic, various immunodetection tests for anti-SARS-CoV-2 IgG and IgM have been suggested. In the systematic review carried out by Lisboa-Bastos and co-workers compared nine ELISA studies to detect SARS-CoV-2 antibodies, they report that the sensitivity of the ELISA on average was 84.3% with a confidence interval (CI 95%) between 75.6% and 90.9% (6). The ELIPSE-Col test has a sensitivity of 91,4%, which places it in the upper limit to that reported in the systematic review. In this same review, the average specificity was 97.6%, ranging between 93.2% and 99.4%, values higher than the 91% found in the present study, but which do not prevent its use in immunodiagnosis, considering the inclusion of negative sera from patients from endemic areas for arboviruses and other circulating coronaviruses in Colombia.

These results constitute an evidence that supports the using of rationaly designed peptides and their strategical mixtures, as important molecular tools useful for detection of antibodies to SARS-CoV-2, revealing acceptable specificity and sensitivity as an immunodiagnostic test.

## Data Availability

All data will be available at the final publication

## FUNDING

This study was funded by Ministerio de Ciencia Tecnología e Innovación (MinCiencias) (project number: 2104101577317, grant number: 361-2020) and by the National Institute of Health, Bogota, Colombia.

## DECLARATION OF COMPETING INTEREST

The authors declare no competing interest.

## ACKNOWLEDGEMENTS

The authors thank the entire team of researchers of the project “Seroprevalence of SARS-CoV-2 during the epidemic in Colombia: a country study” for the joint work with the professionals involved in facing the SARS-CoV-2 pandemic from different aspects and to the Directorates of Public Health Research. The authors thank all the Colombian and foreign researchers who deposited genomes in GISAID’s EpiFlu (TM).

## Notes

### Competing Interest Statement

The authors have declared no competing interest.

### Funding Statement

This study was funded by Ministerio de Ciencia Tecnologia e Innovacion (MinCiencias) (project number: 2104101577317, contract number: 361-2020) and by the National Institute of Health, Bogota, Colombia

### Author Declarations

This work was approved by the Ethics and Research Methodologies Committee (CEMIN) of the National Institute of Health (created by resolution 395 of April 4, 2017) who determined that the work meets the technical and ethical requirements for which they granted ethical approval registered in Act No. 7 of April 13, 2020. This work was developed according to the national law 9/1979, decrees 786/1990 and 2323/2006, which establishes that the Instituto Nacional de Salud (INS) from Colombia is the reference laboratory and health authority of the national network of laboratories and in cases of a public health emergency or those in which scientific research for public health purposes as required. This study was performed following the ethical standards of the Declaration of Helsinki 1964 and its later amendments. The information used for this study comes from secondary sources of data that were previously anonymized and do protect patient data. All necessary patient/participant consent has been obtained and the appropriate institutional forms have been archived.

